# Clinical and electrophysiological effects of cathodal HD-tDCS to the right STS in psychosis spectrum disorders: A pilot proof of concept study

**DOI:** 10.64898/2026.01.21.26344308

**Authors:** Rebekah L Trotti, Nicolas Raymond, David A Parker, Halide Bilge Turkozer, Prachi Patel, Brendan Stiltner, Daphne Ying, Willa Molho, Robert MG Reinhart, Matcheri S Keshavan, Paulo Lizano

## Abstract

The right superior temporal sulcus (rSTS) plays a causal role in multisensory hallucinations, is associated with audiovisual integration (AVI), and displays abnormal activity in psychotic disorders. We sought to reduce psychosis symptoms including hallucinations while modulating AVI and rSTS activity with cathodal, high-definition transcranial direct current stimulation (HD-tDCS) of the rSTS. This double-blind, randomized, sham-controlled pilot study applied HD-tDCS to the rSTS using a ring montage (-1.5 mA cathode center and three .5 mA anode surrounds). Stimulation was administered (N=6 active, N=6 sham) for 5 days with 2 20-minute sessions per day. Assessments occurred at baseline, 5-day, and 1-month timepoints. Electroencephalography (EEG) was recorded during resting state and an AVI steady state (SSR) paradigm. Given the pilot nature of this study, analyses used an alpha threshold of .10. The active group demonstrated reduced clinician-assessed symptoms at 5-days and 1-month, attenuated AVI at 5-days, enhanced auditory SSR and attenuated alpha power over the rSTS at 1-month, and reduced self-reported symptoms at 5-days. Symptom changes correlated with alpha changes. Biological motion, functioning, mood, and cognition did not significantly change. Findings suggest therapeutic effects and successful target engagement. Results provide preliminary proof-of-concept evidence supporting the initiation of a larger trial in psychotic disorders.

## Introduction

Auditory and visual hallucinations are common, prominent symptoms of psychosis spectrum disorders (PSD) like schizophrenia, and their presence predicts disability and disease outcomes^1^. Existing treatments for hallucinations are often poorly tolerated, and up to 30% of patients are treatment resistant^2,3^. Despite decades of neuroscience research, few new or targeted treatments for hallucinations have been established. Transcranial electrical stimulation (tES) is informed by known neurological correlates of psychiatric illness and may provide a novel, well-tolerated treatment for hallucinations. tES modulates neural activity by applying anodal or cathodal direct current to the cortex to affect the underlying neural membrane potential. High-definition direct current stimulation (HD-tDCS) increases spatial precision, allowing researchers to target specific cortical regions.

Though many areas have been implicated in hallucination production, lesion network mapping^4^ provides strong causal evidence for the role of the right superior temporal sulcus (rSTS) in hallucinations^5^, suggesting it may be an ideal tES treatment target. This region is negatively connected with more than 90% of lesions causing hallucinations^5^. Thus, when spontaneous activity decreases at lesion locations causing hallucinations, spontaneous activity in the rSTS increases. Thereby applying cathodal HD-tDCS to the rSTS may affect excitability of this region, modulating network dynamics to reduce the occurrence of hallucinations in people with PSD.

The rSTS is active during multisensory hallucinations, receiving convergent somatosensory, auditory, and visual inputs. It is involved in audio-visual integration (AVI), which is impaired in people with PSD^6–10^ and shares a strong association with hallucination severity^11^. We reasoned that rSTS hyperexcitability may lead to hallucinations and increased global symptom burden by aberrant multisensory integration. AVI may be studied with electroencephalography (EEG) using steady-state stimuli, which are repetitive stimuli presented at a set frequency. Cortical neurons in the relevant sensory network entrain to the stimulus frequency, firing in a time-locked fashion at the stimulus’ driving frequency, and eliciting a steady state response (SSR)^12,13^. When auditory and visual steady-state stimuli are presented simultaneously, AVI may induce additional “intermodulation” frequencies^14,15^. We used this novel approach to measure AVI in this trial.

Beyond its roles in AVI and hallucinations, the STS is involved in numerous other functions and behaviors including social cognition and perception, biological motion, theory of mind, speech and language^16,17^ and general constructs like quality of life^18^.

Due to these roles, rSTS modulation may also have a broader therapeutic effect on symptoms of PSDs. This trial will collect data on a range of psychiatric symptoms and behaviors to track these and other potential effects of neuromodulation.

We conceptualized hallucinations in PSD as arising from aberrant multisensory binding within the rSTS, reflected in exaggerated intermodulation responses and altered intrinsic oscillatory activity. Cathodal HD-tDCS may normalize this activity, producing both neurophysiological and clinical change. To examine this hypothesis, this double-blind, sham-controlled pilot study examined if a 5-day course of twice daily, 20-minute cathodal HD-tDCS of the rSTS compared to sham stimulation may reduce clinical indicators of psychosis and hallucinations and enhance brain activity. We hypothesized that hallucinations will decrease and intermodulation frequency power will change from baseline to 5 day and 1-month follow-up in the active tDCS group but not sham. Given our directional hypothesis, we used a liberal alpha (α=.10) in this pilot proof of concept study. We also conducted exploratory analyses investigating if other psychosis symptoms, biological motion detection, global function, mood, and cognition change with stimulation.

## Methods

### Participants

This trial employed a randomized, double blind, parallel-arm design and took place at Beth Israel Deaconess Medical Center in Boston, MA. Inclusion criteria comprised diagnosis of schizophrenia, schizoaffective disorder, or psychotic bipolar disorder by the Structured Clinical Interview for DSM-V^19^, lifetime history of hallucinations, 18-80 years old, no changes to relevant antipsychotic medications for 1 month prior to participation, and a sufficient level of English for participation. Exclusion criteria included pregnancy or breastfeeding, IQ <60, any major neurologic illness, substance use disorder within 6 months of participation, positive urine drug screen, a history of moderate-to-severe visual impairment, significant suicidal ideation within 6 months of participation, serious medical illness or instability requiring hospitalization within the past year, metallic or electronic implants, recent or serious cardiac events, and skin sensitivity or claustrophobia precluding them from completing study protocols.

12 individuals with psychosis were enrolled and randomized to either sham (n=6) or active HD-tDCS (n=6; Figure 1). 12 participants completed a 5-day follow-up and 11 completed a 1-month follow-up. Participants and researchers involved in data collection and analysis were blind to study assignment.

**Figure 1.**
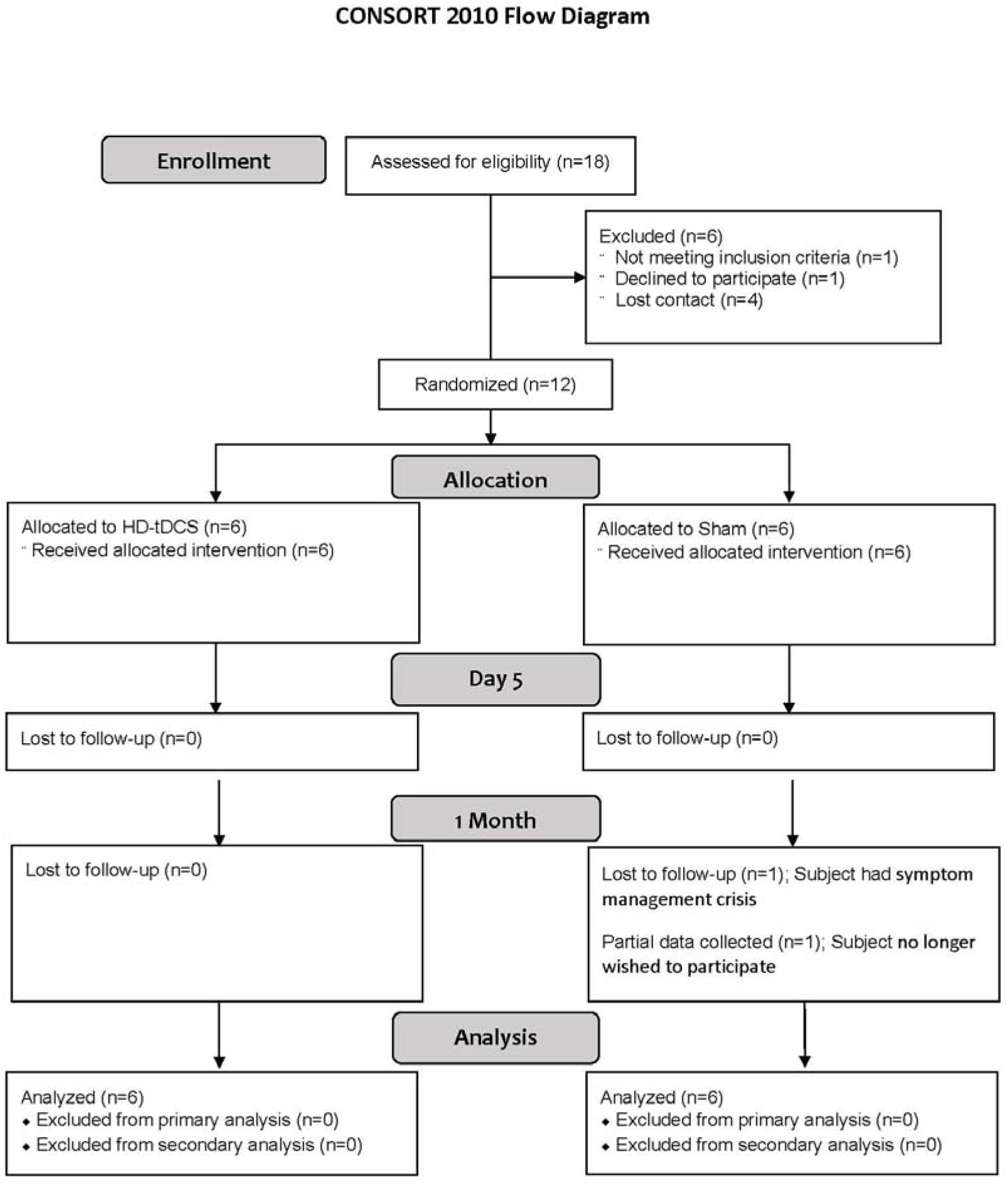
CONSORT diagram for recruited participants in this trial.

All participants provided written informed consent prior to participation after obtaining a complete description of study procedures. The study was approved by the Institutional Review Board at Beth Israel Deaconess Medical Center (2019P0001016).

### Procedures

HD-tDCS was administered using an MxN 9-channel HD-tES system (Soterix Medical) with sintered 12 mm Ag/AgCl electrodes in plastic holders, filled with conductive gel, and embedded in the Quik-Cap. Montage selection was guided by current flow modeling in HD-Explore (Soterix Medical) and ROAST^20^, with the chosen montage balancing high focality and intensity at MNI coordinate x=61, y=-20, z=-3. This montage includes one cathode (-1.5mA) at T8, surrounded by 3 anodes (.50mA each) at FT8, C6, and TP8 (Figure 2).

**Figure 2.**
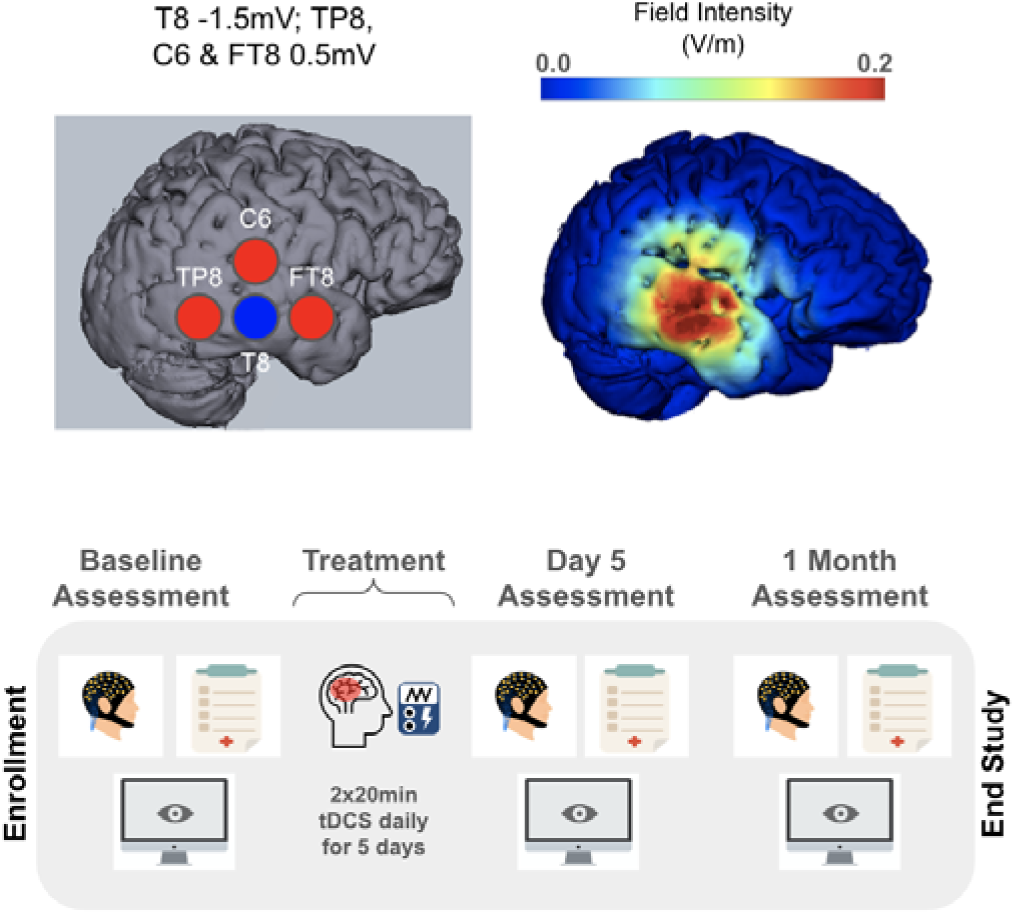
Study methods. The stimulation electrodes are shown at the top left with the cathode in blue and anodes in red. The resulting current flow model is shown at the top right, focused on the rSTS. The study timeline is shown on the bottom, depicting a baseline assessment, 5-day course of daily tDCS, 5-day follow-up, and 1-month follow-up.

Participants received 2 consecutive 20-minute HD-tDCS sessions separated by a 5-10 minute break on 5 consecutive weekdays (Figure 2), as recommended by Kim et al., 2019^21^. The sham condition used the same montage, with only a 30 second ramp up, 30 second ramp down, and no stimulation for 20-minutes in-between.

Randomization used the ‘blockrand’ package in R Studio^22^ with block size set to 2. An unblinded researcher set the tES device to active or sham for each session out of view from the participant and blinded assessors. Topocaine 5 was applied 5 minutes before stimulation to minimize sensation differences between active and sham. After each session, participants completed a Sensation Questionnaire to gauge blinding adequacy and side effects (see Supplement).

### Primary outcomes

Participants completed the Positive and Negative Syndrome Scale for Schizophrenia (PANSS^23^) by clinician interview at all timepoints and subdomain scores were calculated (Positive, Negative, General).

#### Active hallucinators

Participants experiencing active hallucinations completed the University of Miami Parkinson’s Disease Hallucinations Questionnaire (UM-PDHQ^24^), which measures the severity of any sensory hallucination, and Auditory Hallucination Rating Scale (AHRS^25,26^), which measures voice hearing hallucinations. 6 participants were experiencing active hallucinations (active N=3, sham N=3) and 4 were experiencing voice hearing (active N=1, sham N=3) at baseline. UM-PDHQ analyses used an average score from quantitative data across sensory modalities to simplify comparisons.

### Secondary outcomes: EEG

12 participants completed EEGs at baseline and 5-day follow-up, and 10 completed EEGs at 1-month follow-up. 1 participant was excluded from all timepoints due to poor data quality and 1 was lost to follow-up. EEG employed a 128-channel HydroCel Geodesic Sensor Net (Magstim EGI) with participants in a sound- and light-shielded booth and impedances kept below 50 kΩ. Stimuli were presented using custom code (Neurobehavioral Systems).

#### AVI task

The AVI task contained auditory and visual steady state stimuli that were amplitude- and luminance-modulated at defined frequencies (ASSR: 40 and 80 hz, VSSR central visual field: 15 hz, VSSR peripherals: 12 hz; Supplement Figure S1). 20 trials were auditory-only (600ms; half at 40 hz, half at 80 hz), 20 were visual-only (2000 ms), and 80 were audiovisual stimuli together. Participants were instructed to passively observe stimuli.

#### AVI variable extraction

Data were cleaned using the Harvard Automated Preprocessing Pipeline for Electroencephalography (HAPPE^27^, parameters in Supplement), converted to time-frequency space in Matlab (EEGLab^28^ newtimef), and averaged over trials. Power at fundamental frequencies and their harmonics (VSSR: 12, 15, 24, 30 hz; ASSR: 40 hz) were extracted from sensors over the relevant cortices (Supplement Figure S2) and intermodulation frequencies (55, 52, 70, 64, 25, 10, 28, and 16 hz) were extracted from sensors capturing rSTS activity (Figure S2). To retain one composite measure for each stimulation type, we averaged over fundamental and harmonic frequencies for the VSSR and all intermodulation frequencies for AVI.

#### Resting state EEG

A 5-minute resting state (rsEEG) was collected as participants sat with their eyes loosely fixed on a white plus sign on a black screen. After preprocessing, data were transformed to time-frequency space in EEGLab^28^ (see Supplement) and averaged into defined frequency bands consistent with Thomas et al., 2019^29^: delta/theta (1-8 hz), alpha (8-12 hz), beta (13-30 hz), and gamma (30-55 hz). Power was averaged across sensors for an overall power metric and at stimulation sensors over the rSTS (Figure S2) for a ROI power metric.

### Secondary outcomes: Behavioral and clinical

A biological motion task^30^ was administered where participants indicated which direction a figure was walking, with 3 increasing levels of nuisance dots (noise), to increase difficulty (easy, medium, and hard; 20 trials each). The percentage of correct responses at each level was calculated for analysis (see Supplement).

The Global Assessment of Function (GAF; Axis 5 on the DSM-IV^31^), Montgomery-Asberg Depression Rating Scale (MADRS^32^), and Young Mania Rating Scale (YMRS^33^) were completed by clinician assessment. Cognitive assessment used the Brief Assessment of Cognition (BAC App^34,35^), administered by iPad (VeraSci, WCG Clinical). Composite and subdomain scores (see Supplement) were z-scored and adjusted based on normative scores^35^.

Finally, participants completed the Symptom Checklist-90 Revised (SCL-90R^36^) by self-report and subscores were calculated (Somatization, OCD, Interpersonal, Depression, Anxiety, Phobic, Paranoid, Psychoticism).

### Data analysis

*Primary and secondary endpoints.* Data were compared between baseline, 5-day, and 1-month timepoints. Primary endpoints included PANSS, UM-PDHQ, and AHRS scores. Secondary endpoints included EEG measurements from the AVI task (ASSR, VSSR, AVI) and resting state (4 frequency bands, ROI), biological motion accuracy, and GAF, MADRS, YMRS, BAC, and SCL-90R scores.

#### Statistics

Intent-to-treat analyses were conducted using custom code in R Studio. Multiple imputation was performed for missing datapoints using the “Mice” package^37^ (0% baseline, 4.75% 5-day, 14.04% 1-month; see Supplement). Contrasts employed nonparametric Wilcoxon Signed-Rank tests (within-subjects) and Mann-Whitney U tests (between-subjects). We tested associations between changes in EEG and symptom variables by conducting Spearman correlations on change scores. We created change scores (follow-up minus baseline) for variables that significantly differed between timepoints. Holm-Bonferroni corrections were applied to multiple comparisons and rank biserial effect sizes were calculated. Since there were very few participants with active hallucinations (3 per group at baseline and 1-month, 2 per group at 5-days), only descriptive statistics for the UM-PDHQ and AHRS were calculated.

#### Power analyses

Our prior trial of lesion-network guided tDCS demonstrated within-subjects effect sizes of .87 for clinical outcomes and .97-2.35 for EEG outcomes. Based on these effect sizes, we determined an alpha threshold of .10 would offer adequate power for this study (73% power for clinical and 63-99% power for EEG outcomes). Given that the intent of a pilot trial is signal detection, this threshold also has the advantage of minimizing false negatives. Following analysis, we used GPower^38^ to complete post-hoc power analyses to estimate achieved power and a priori analyses to estimate appropriate sample sizes for a future confirmatory efficacy trial, reported in the Supplement (Table S12).

## Results

### Demographics

Individuals were diagnosed with schizophrenia (N=2), schizoaffective disorder (N=5), and bipolar I disorder with psychosis (N=5). Sex distribution was equal between the sham and active groups (50% female). Age did not differ between sham (M=37.5, SD=14.84) and active (M=36.67, SD=8.26; *F*(1)=.01, *p*=.90). Groups also did not significantly differ on outcome variables at baseline (all *p*>.10).

### Primary endpoints

While few results survived corrections for multiple comparisons, statistical tests revealed several significant effects that suggest feasibility and mechanistic engagement.

#### PANSS

In the active group, PANSS Total significantly decreased from baseline to 5-days (p=.03, p_adj=_.09) and 1-month (p=.05, p_adj_=.11; Figure 3). Positive scores also decreased at 5-days (p=.09, p_adj_=.27) and General scores decreased between baseline and 1-month (p=.08, p_adj_=.26). There were no significant changes in the Negative subscale or sham group (full statistics and effect sizes in Table 1).

**Figure 3.**
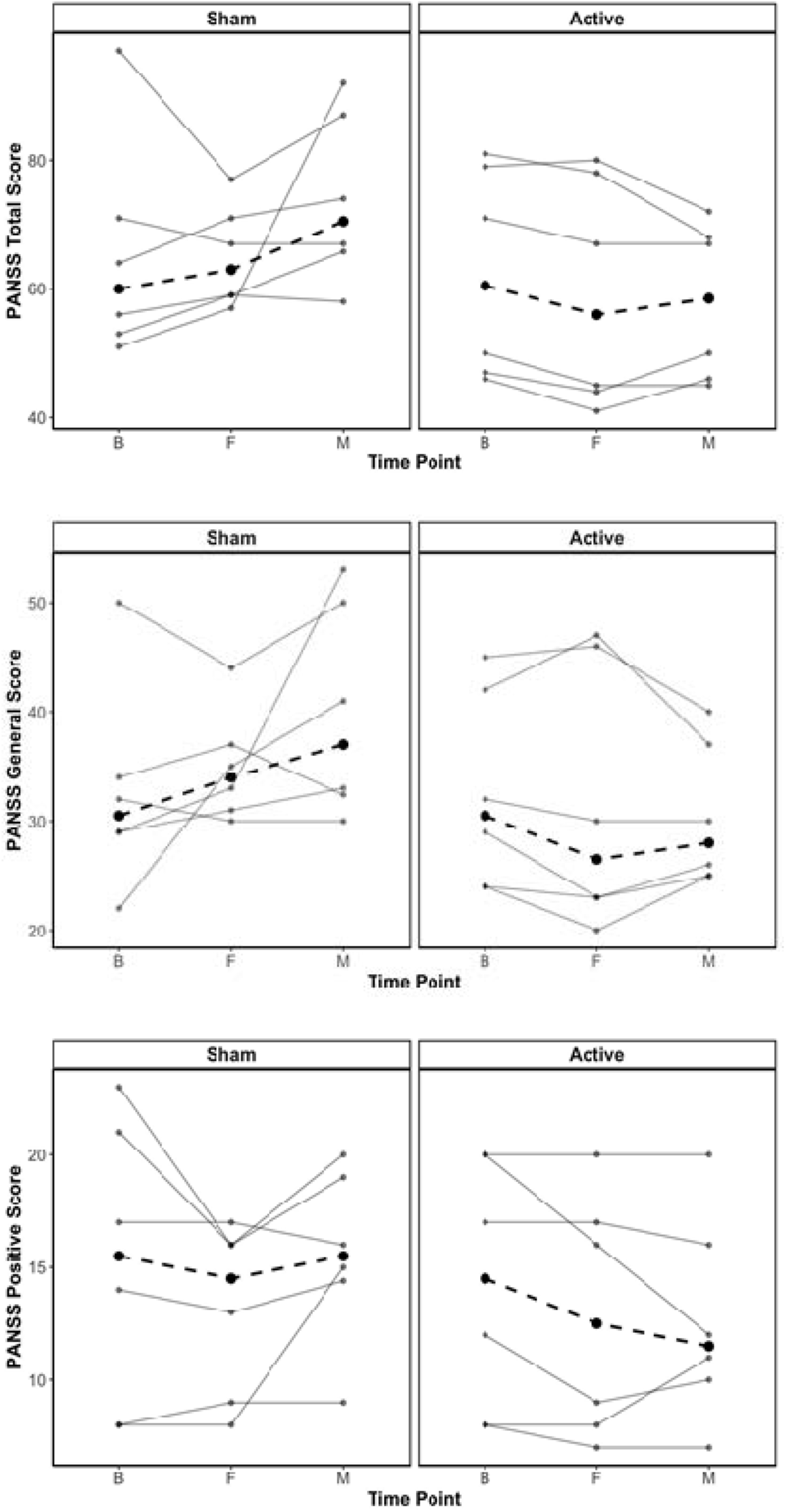
PANSS Total, Positive, and General scores for participants in the sham and active groups at baseline, 5-days, and 1-month (B/F/M). Solid lines represent individual participant data and bold dotted lines represent the group average.

**Table 1.** Positive and Negative Syndrome Scale Results.

|  | Comparison | Statistic | p | p adjusted | Rank Biserial r |
| --- | --- | --- | --- | --- | --- |
| Active HD-tDCS |  |  |  |  |  |
| Total | B vs F | 20 | **0.03\*** | **0.09\*** | 0.90 |
|  | B vs M | 14 | **0.05\*** | 0.11 | 0.87 |
|  | F vs M | 7 | 0.29 | 0.29 | 0.40 |
| Positive | B vs F | 6 | **0.09\*** | 0.27 | 1.00 |
|  | B vs M | 11 | 0.21 | 0.42 | 0.47 |
|  | F vs M | 5.5 | 0.50 | 0.50 | 0.10 |
| Negative | B vs F | 6.5 | 0.36 | 0.71 | 0.30 |
|  | B vs M | 8.5 | 0.13 | 0.40 | 0.70 |
|  | F vs M | 3 | 0.61 | 0.71 | 0.00 |
| General | B vs F | 14.5 | 0.23 | 0.46 | 0.38 |
|  | B vs M | 17.5 | **0.09\*** | 0.26 | 0.67 |
|  | F vs M | 9 | 0.39 | 0.46 | 0.20 |
| Sham HD-tDCS |  |  |  |  |  |
| Total | B vs F | 8 | 0.74 | 1.00 | -0.24 |
|  | B vs M | 5.5 | 0.88 | 1.00 | -0.48 |
|  | F vs M | 1 | 0.97 | 1.00 | -0.87 |
| Positive | B vs F | 8.5 | 0.13 | 0.40 | 0.70 |
|  | B vs M | 11.5 | 0.46 | 0.92 | 0.10 |
|  | F vs M | 1 | 0.97 | 0.97 | -0.87 |
| Negative | B vs F | 11 | 0.50 | 1.00 | 0.05 |
|  | B vs M | 6 | 0.71 | 1.00 | -0.20 |
|  | F vs M | 5 | 0.79 | 1.00 | -0.33 |
| General | B vs F | 6.5 | 0.83 | 1.00 | -0.38 |
|  | B vs M | 3 | 0.91 | 1.00 | -0.60 |
|  | F vs M | 2 | 0.95 | 1.00 | -0.73 |

*Note.* Statistics were performed with one-tailed Wilcoxon Signed-Rank Tests

#### Active hallucinations

One participant per group stopped experiencing hallucinations at 5-days and one participant in the active group started experiencing low-intensity hallucinations at 1-month. The average UM-PDHQ score decreased for two active participants between baseline and 5-day and remained stable at 1-month. Sham participants remained approximately the same across time (Figure 4).

**Figure 4.**
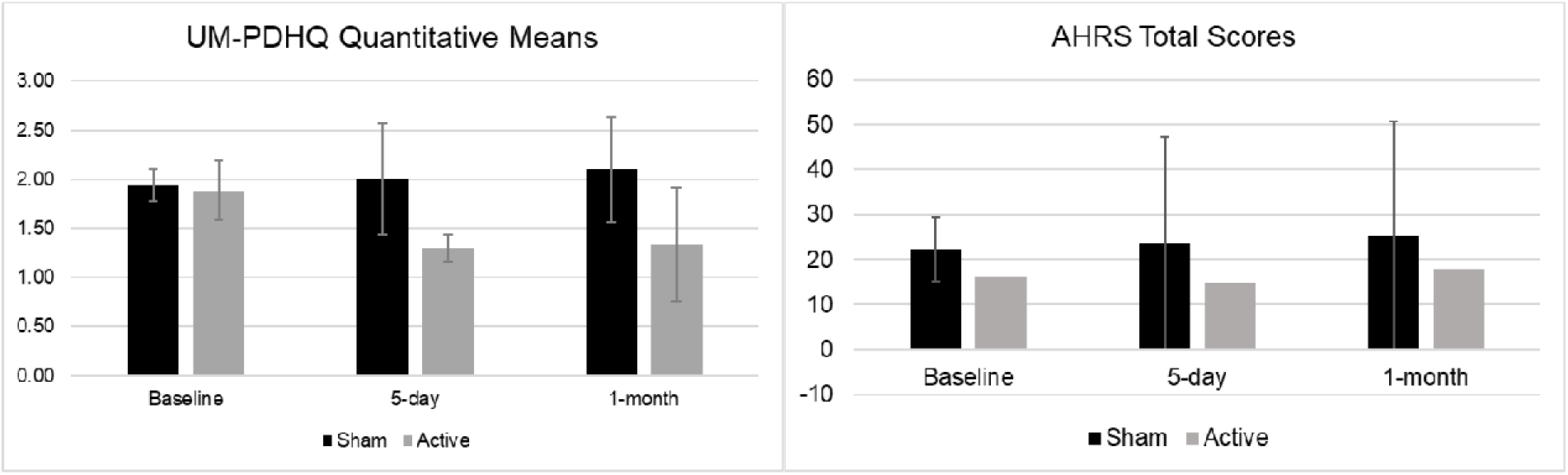
Active hallucination descriptive statistics. The average score for active tDCS participants (N=3) decreased at 5 day and 1 month but remained stable for sham participants (N=3). For the AHRS, the active participant’s score decreased by 1 while the sham average score slightly increased (N=3).

For the AHRS, the active participant experienced a 1-point increase in voice hearing frequency, but a 1-point decrease in how real they seem and loudness at the 5-day timepoint, for a total decrease of 1 point. At 1-month, all scores returned to baseline, but voice extensiveness increased by 2 points. In the sham group, AHRS Total slightly increased for 2 out of 3 participants at each timepoint (Figure 4).

### Secondary endpoints

#### Audiovisual EEG

In the active group, ASSR significantly increased from 5-days to 1-month (p=.06, p_adj_=.18) but did not change between baseline and 5-days (p=.79; Figure 5). The active group also experienced a significant decrease in AVI from baseline to 5-days (p=.06, p_adj_=.18; Figure 5), but no change at 1-month (p>.10). VSSR did not significantly change, and there were no significant effects in sham (all p>.10; Table 2).

**Figure 5.**
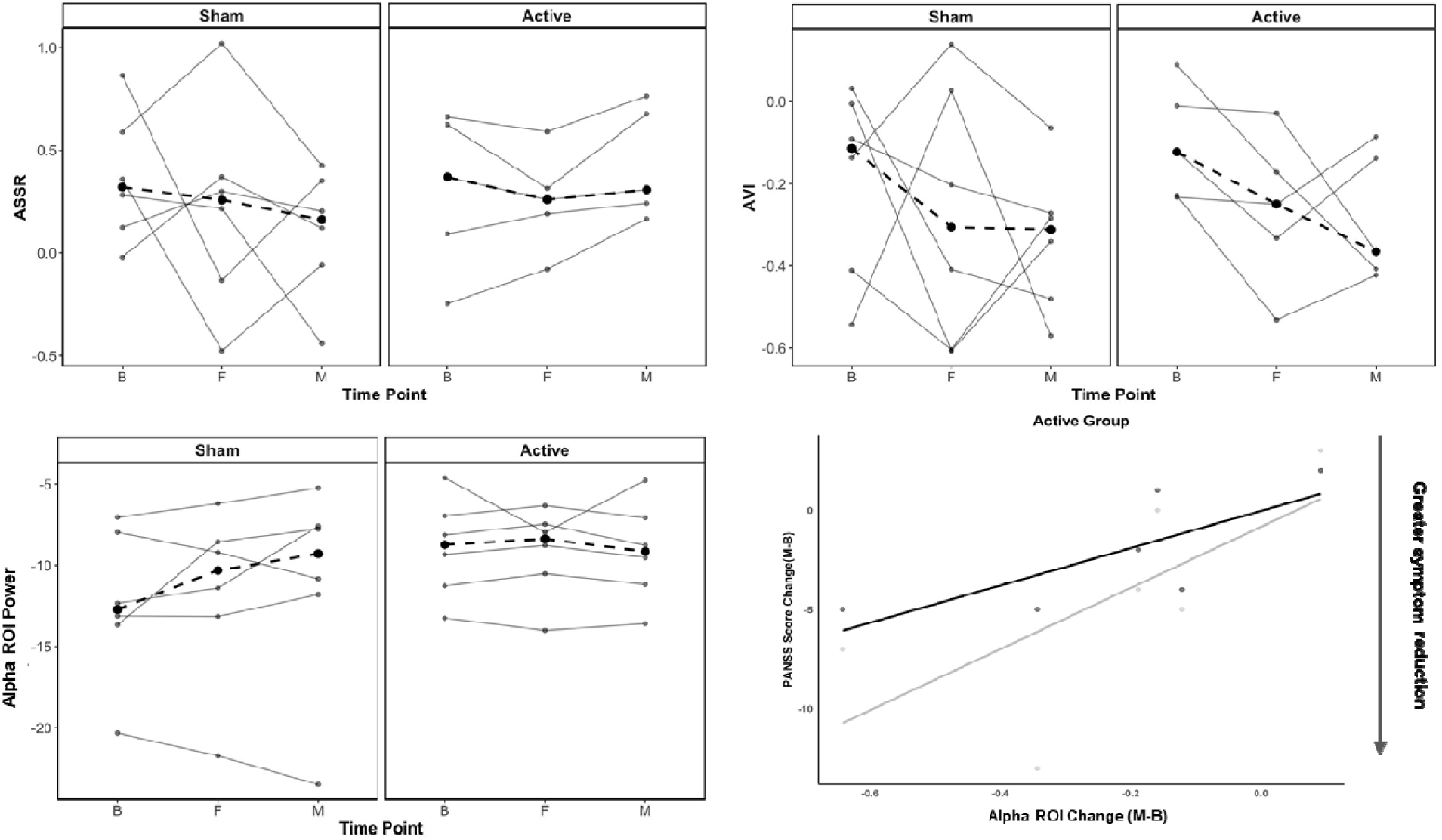
rsEEG, ASSR, and AVI scores and correlations at baseline, 5-day, and 1-month (B/F/M). There were significant effects over time in the active group, but not in the sham group. Solid lines represent individual participant data and bold dotted lines represent the group average. Alpha ROI change correlated with changes in both PANSS Total and General.

**Table 2.** Audiovisual EEG Results.

|  | Comparison | Statistic | p | p adjusted | Rank Biserial r |
| --- | --- | --- | --- | --- | --- |
| Active HD-tDCS |  |  |  |  |  |
| ASSR | B vs F | 9 | 0.79 | 0.79 | 0.20 |
|  | B vs M | 2 | 0.18 | 0.36 | -0.73 |
|  | F vs M | 0 | **0.06\*** | 0.18 | -1.00 |
| AVI | B vs F | 15 | **0.06\*** | 0.18 | 1.00 |
|  | B vs M | 13 | 0.18 | 0.36 | 0.73 |
|  | F vs M | 9 | 0.79 | 0.79 | 0.20 |
| VSSR | B vs F | 2 | 0.18 | 0.36 | -0.73 |
|  | B vs M | 1 | 0.11 | 0.32 | -0.87 |
|  | F vs M | 6 | 0.79 | 0.79 | -0.20 |
| Sham HD-tDCS |  |  |  |  |  |
| ASSR | B vs F | 12 | 0.83 | 1.00 | 0.14 |
|  | B vs M | 18 | 0.14 | 0.43 | 0.71 |
|  | F vs M | 14 | 0.53 | 1.00 | 0.33 |
| AVI | B vs F | 13 | 0.67 | 1.00 | 0.24 |
|  | B vs M | 16 | 0.29 | 0.88 | 0.52 |
|  | F vs M | 12 | 0.83 | 1.00 | 0.14 |
| VSSR | B vs F | 11 | 1.00 | 1.00 | 0.05 |
|  | B vs M | 18 | 0.14 | 0.43 | 0.71 |
|  | F vs M | 13 | 0.67 | 1.00 | 0.24 |

*Note.* Statistics were performed with one-tailed Wilcoxon Signed-Rank Tests. ASSR=Auditory Steady-State Response, AVI=Audiovisual Integration, VSSR=Visual Steady-State Response

#### Resting EEG

In the active group, alpha power over the rSTS significantly decreased between baseline and 1-month (p=.06, p_adj=_.18), but not at 5-days (p=1.0; Figure 5). All other measures and frequencies did not significantly change, and there were no effects in sham (all p>.10; Table 3).

**Table 3.**
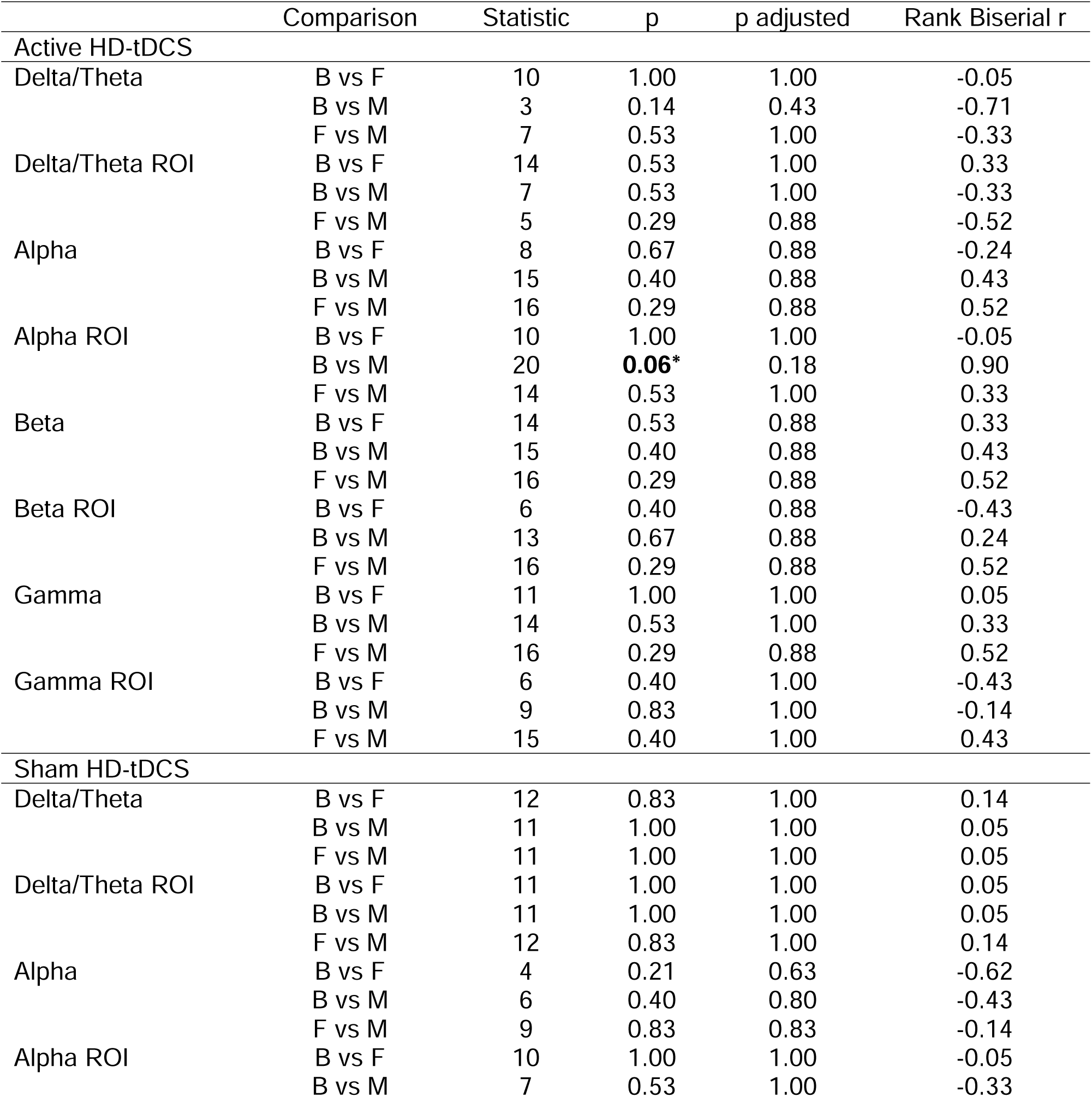
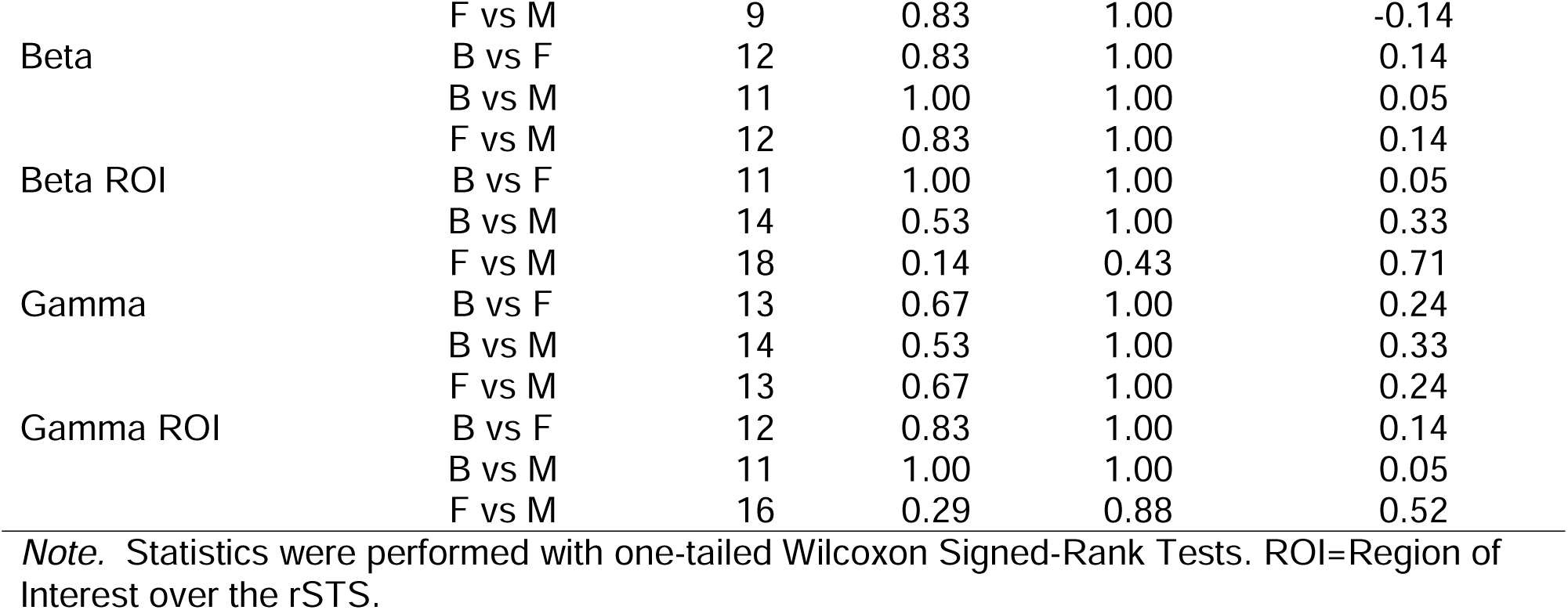
Resting State EEG Results.

#### Off-target effects

There were no significant changes in biological motion for either group (all p>.10). In sham, YMRS total and BAC Composite significantly decreased from Baseline to 1-month (YMRS: p=.06, p_adj_=.18, BAC: p=.04, p_adj_=.11). There were no significant changes in the active group or changes in the GAF, MADRS, or BAC subscales (all p>.10; Supplement Tables 3-4).

#### Self-reported symptoms

In the active group, SCL90-R Total significantly decreased at 5-days (p=.06, p_adj_=.18), explained by reductions in Somatization, OCD, and Anxiety (Figure 6, Table S5). The active group’s SCL90-R Total returned to baseline at 1-month (p=.01, p_adj_=.11), explained by significant increases in Somatization and Interpersonal scores (Figure 6, Table S5). In sham, SCL90-R Total significantly increased from baseline to 5-days (p=.06, p_adj_=.18) and Somatization significantly decreased from baseline to 1-month (p=.09, p_adj_ = .19) and 5-days to 1-month (p=.01, p_adj_=.11; all statistics in Supplement Table S5).

**Figure 6.**
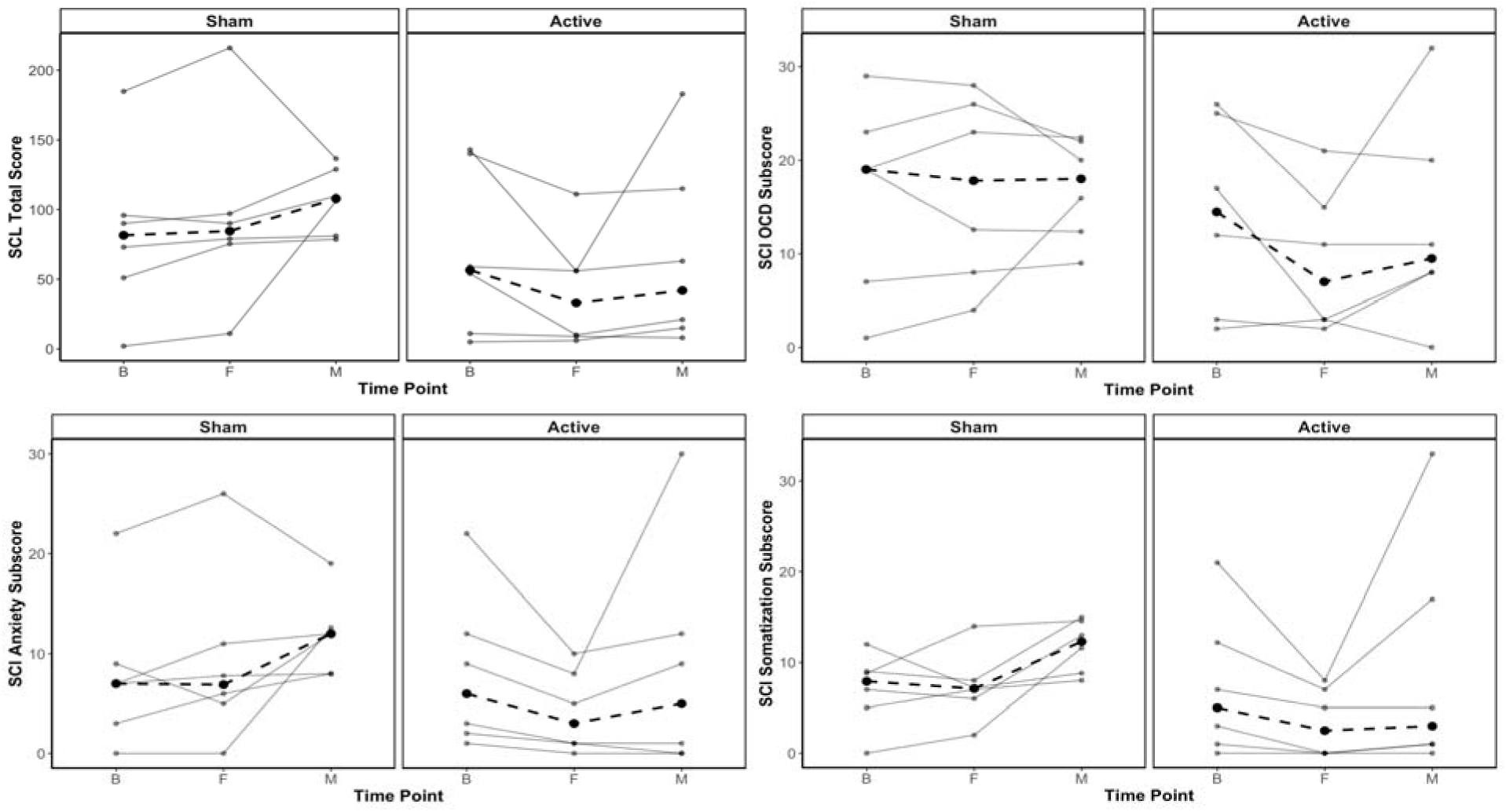
Changes in the SCL-90 between baseline, 5-day, and 1-month timepoints (B/F/M). Solid lines represent individual participant data and bold dotted lines represent the group average.

### Correlations

Change scores were calculated by subtracting Baseline values from both 5-day and 1-month data for measures that significantly changed in the active group (clinical: PANSS Total, Positive, General, SCL90-R; EEG: alpha ROI rsEEG, ASSR, AVI), resulting in 6 total correlations. 1-month change in alpha ROI rsEEG was significantly and positively correlated with PANSS Total (p=.07, p_adj_=.35, r=.77) and PANSS General changes (p=.05, p_adj_=.30, r=.81; Figure 5).

## Discussion

This pilot HD-tDCS trial examined if cathodal rSTS stimulation could reduce psychosis symptoms, including hallucinations, and improve audiovisual integration. Though this was a small proof-of-concept trial and few p-values survived corrections, results generally supported hypotheses and merit further study. In the active group, symptoms improved by clinician and self-report along with a qualitative decrease in hallucination measures, indicating preliminary clinical efficacy. EEG measures showed modulation of auditory processing, audiovisual integration, and alpha activity over the stimulated cortex, suggesting successful engagement of the rSTS. Further, PANSS and alpha changes significantly correlated, indicating a mechanistic link between neurophysiology and symptoms. Overall, this trial provided preliminary evidence that cathodal stimulation could affect rSTS network dynamics to reduce psychosis, hallucinations, and other symptoms of psychotic disorders.

Our primary hypothesis was that cathodal rSTS stimulation would reduce hallucinations given the causal role of the rSTS in hallucination production, which was partially supported by results. PANSS Positive decreased at 5-days and trend-level results suggested reduced UM-PDHQ and AHRS-measured hallucinations that were preserved at 1-month. Though hallucination analyses were underpowered, they are supported by our prior case study, which showed a substantial drop in AHRS with rSTS stimulation^39^ and changes in ASSR, suggesting a specific impact of stimulation on sensory processing. In future trials, we will continue to examine the effects of stimulation on hallucinations, given their prevalence in psychotic disorders.

The potential therapeutic effect of stimulation was best reflected in the PANSS Total and General, and supported by the SCL90-R, indicating that both clinicians and patients can detect clinical improvement following HD-tDCS. PANSS Total scores decreased with a large effect at 5-days that was preserved at 1-month, indicating a short-term effect of stimulation that plateaus over time. PANSS General also decreased at 1-month, suggesting broad effects on symptoms other than hallucinations.

Additionally, SCL90-R supported these effects, with the active group’s Total scores decreasing at 5-days, accounted for by reduced somatization, OCD, and anxiety. As the STS is a core part of networks underlying numerous functions like social skills, face processing, speech processing, and theory of mind^40^, this range of potential clinical effects could be due to stimulation’s modulation of these STS networks. Other noninvasive brain stimulation studies targeting the STS have similarly modulated face recognition^41^, relationship memory^42^ and biological motion^43^. Downstream effects of these basic processes, like social functioning and complex cognition, may explain broad effects of HD-tDCS on psychiatric symptoms. Subsequent studies with increased statistical power and additional measures will further examine these possibilities.

Physiologically, we used audiovisual and resting state EEG to examine engagement of the rSTS. Though the directionality of neurophysiological AVI deficits in psychosis is unclear^44^, some studies suggest that neural hyperactivity during multisensory facilitation may characterize psychosis spectrum disorders^45^. This prior study would suggest that our finding of reduced AVI power at 5-days could indicate neurophysiological improvement, where HD-tDCS moved neural activity closer to healthy levels. Similarly, there was a decrease in alpha rsEEG power over the rSTS at 1-month that correlated with PANSS changes, reinforcing the hypothesis that stimulation successfully engaged this region, normalized its activity, and is associated with behavioral changes, though directional causality is still unknown.

Though results are preliminary, enhanced ASSR, reduced AVI, reduced rSTS alpha power, and a correlation with behavior suggest that stimulation has an effect on its targeted cortex, consistent with our hypothesis that HD-tDCS will modulate rSTS activity to induce clinical improvement. Future studies will further assess these neurophysiological changes, including source analysis, behaviorally-relevant sensory tasks, and causal statistics, to test potential mechanisms underlying effects.

Regarding off-target effects, no within-group changes were seen in mood, functioning, or cognitive measures for the active group. Effects of sham on YMRS and BAC could be explained by practice effects, low power, or regression to the mean.

Additionally of-note, the active group also did not demonstrate full stability on biological motion (see Supplement), suggesting that active stimulation may have slightly modified this function given the proximity of the rSTS to the motion processing region, V5/MT. In future studies, we will consider personalizing stimulation to individual neuroanatomy to more precisely target the intended region.

Strengths of our study include its novelty and a mechanistic approach, but the small sample size is a limitation. Results of this pilot study are promising, suggesting that cathodal rSTS stimulation is potentially efficacious for psychosis symptoms and successfully engages rSTS neurophysiology. Also notably, no significant adverse effects were reported (Supplement Figure S3), indicating that this novel therapy is feasible and safe. Larger clinical trials are needed to better evaluate suitability of the approach for clinical practice and should consider samples enriched with hallucinator participants, personalized neuroanatomical targeting, and pre-specified EEG biomarkers.

## Data Availability

All data produced in the present study are available upon reasonable request to the authors

