## Supplementary Material for "Clinical and electrophysiological effects of cathodal HD-tDCS to the right STS in psychosis spectrum disorders: A pilot proof of concept study"

**Contents:**

1. EEG Preprocessing
2. Figure S1: AVI EEG task
3. Figure S2: Plot of EEG sensor map depicting clusters
4. Biological Motion
5. Table S1: Biological Motion CCC Results
6. Table S2: Biological Motion Wilcoxon Results
7. Table S3: GAF, MADRS, & YMRS Results
8. BAC Subdomains
9. Table S4: BAC Results
10. Table S5: Symptom Checklist 90 Results
11. Imputation
12. Table S6: Unimputed PANSS Results
13. Table S7: Unimputed AVI EEG Results
14. Table S8: Unimputed rsEEG Results
15. Table S9: Unimputed Biological Motion Results
16. Table S10: Unimputed GAF, MADRS, & YMRS Results
17. Table S11: Unimputed BAC Results
18. Power Analyses
19. Table S12: Power Analyses
20. Sensation Questionnaire Results

**EEG Preprocessing**

AVI data were bandpass filtered from .5 to 100 hz and eye blink, cardiac, and muscle artifacts were minimized using ICA. Data were segmented from 1250 ms pre-stimulus to 3850 ms post-stimulus and downsampled to 500 hz. Channels were interpolated within segments, with trials containing ±120 µV amplitude after interpolation at any sensor rejected. Data with <75% of trials per condition included in each subject’s waveform averages were excluded from analysis. After conversion to time-frequency space, power values were converted to decibels, baseline-adjusted using a 150 ms pre-stimulus period, and averaged over sensor clusters (VSSR: 72, 71, 76, 70/O1, 75/Oz, 83/O2, 74, 82, 81; ASSR: 19, 11/Fz, 4, 20, 12, 5, 118, 30, 13, 6, 112, 105, 7, 106, Cz).

Resting state data were bandpass filtered from .5 to 55 hz with a notch filter at 60 hz, and eye blink, cardiac, and muscle artifacts were minimized using Independent Component Analysis (ICA). Data were segmented into 5-second epochs and downsampled to 500 hz. Channels were interpolated within segments. Trials with ±120 µV amplitude at any sensor were rejected. Data with <30 epochs were excluded from analysis. Data were converted to time-frequency space by conducting Fast Fourier Transforms (FFTs) on Hanning tapered windows (500 ms steps, 1 Hz resolution) for each epoch. Power values (squared absolute values of complex FFT outputs) were then converted to decibels (10*log10) and averaged over time.

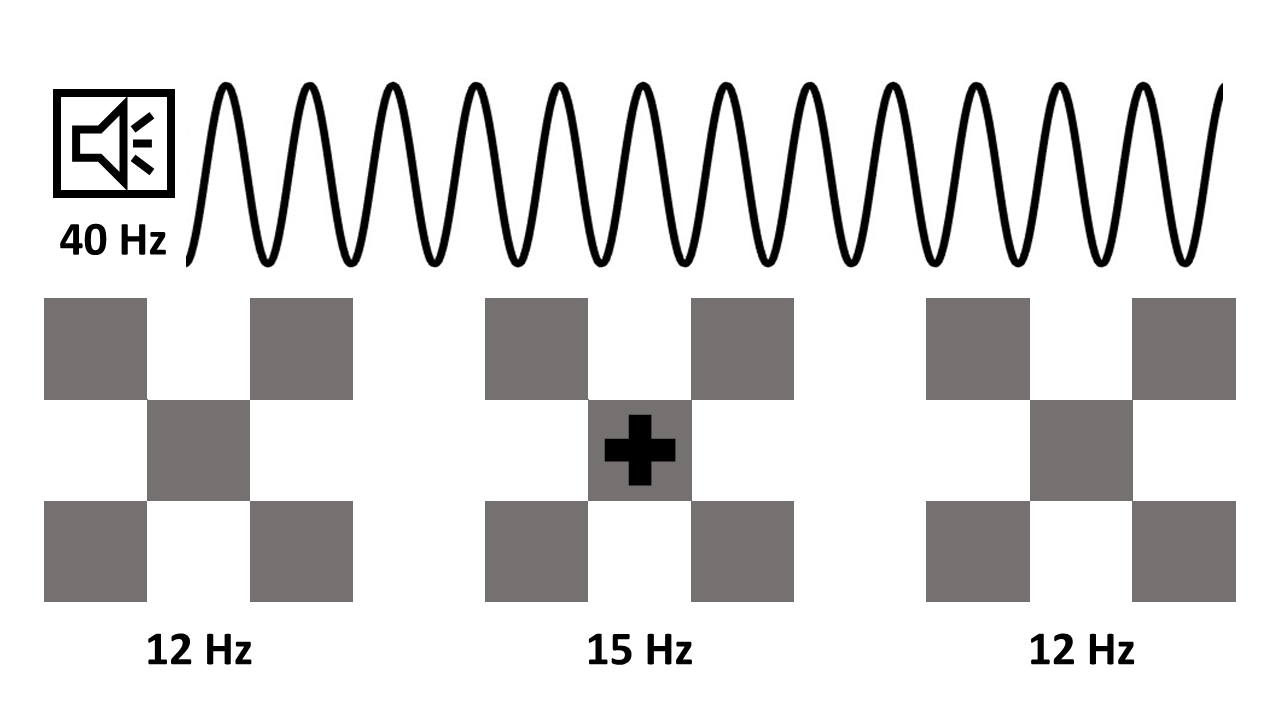
**Figure S1.** AVI EEG Task. Auditory stimulus amplitude was modulated at 40 hz and delivered through headphones. Visual stimuli were grey and white checkerboards to the left, right, and center of a computer screen. Peripheral checkerboards were luminance modulated at 12 hz and the center checkerboard was modulated at 15 hz. Each stimulus was presented alone in ASSR-only and VSSR-only trials and the 2 stimulus types were presented simultaneously in AVI trials

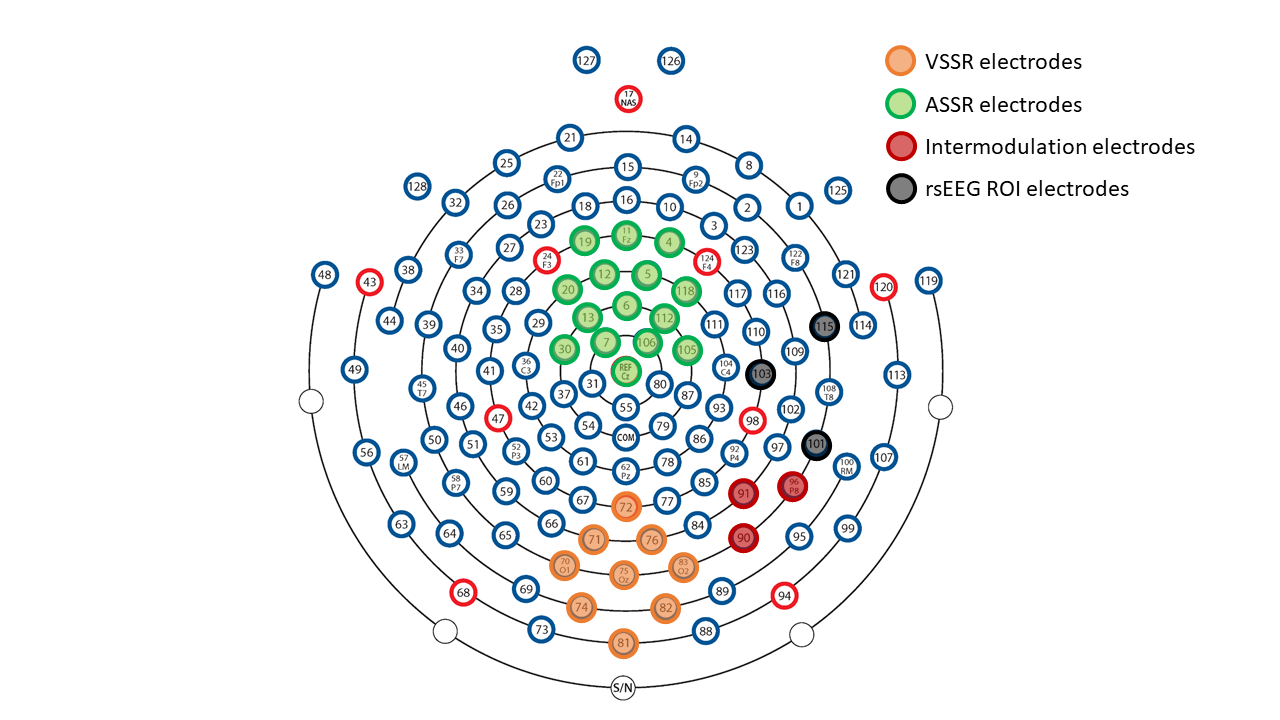

**Figure S2.** Plot of EEG sensor map depicting sensor clusters used in the analyses. The cluster in orange was used to quantify the visual steady state (VSSR) and the cluster in green was used to quantify the auditory steady state (ASSR) at driving frequencies (15 hz, 40 hz). The cluster in red, capturing activity from the rSTS, was used to quantify intermodulation frequencies. The cluster in grey line up with stimulation sensors and were used as an ROI for resting state analyses.

**Biological Motion**

In addition to hallucinations and audiovisual integration, a posterior region of the STS is also involved in processing biological motion (Puce & Perrett, 2003). While we intended to focally target the specific region associated with hallucinations (MNI coordinates c=61, y=-20, z=-3), we tested targeting accuracy and focality by incorporating a point-light biological motion task (Vonck et al., 2015) evaluating participants’ ability to recognize the directional motion of a walking body with visual noise. The point-light figure presented was made of dots with legs, arms, and a head. Tasks were administered on the same computer monitor to all participants at a distance of approximately 100 cm. Lights were turned off, and participants used a chin rest and responded using arrow-keys. Auditory feedback was provided if participants responded correctly or incorrectly in the form of two differently pitched tones; participants were informed which tone indicated a correct and incorrect response. A practice trial without nuisance dots was administered to familiarize participants with the task and assess their understanding of the instructions.

To statistically assess the stability of biological motion, consistent with the hypothesis that this remained unchanged due to stimulation focality, we employed Lin’s Concordance Correlation Coefficient (CCC), the nonparametric equivalent of an Intraclass Correlation Coefficient (ICC). In this test, a significant result indicates that the measure is stable over time. This test is most appropriate to evaluate the hypothesis that biological motion detection does not change with rSTS stimulation.

CCCs did not indicate full stability within the active stimulation group. In the active group, accuracy was only significantly stable at the medium level from baseline to 5-days (CCC=.79 [.37, .94]), whereas in the sham group, accuracy was significantly stable at the easiest level from baseline to 5-days and baseline to 1-month (CCC=.73 [.3, .91]), the medium level across all timepoints (CCCs=.71-.79), and the hardest level from baseline to 5-days (CCC=.75 [.17, .94]; all statistics in Table S1).

| **Table S1.** Biological Motion Concordance Correlation Coefficient Results | | | | | | | | | |
| --- | --- | --- | --- | --- | --- | --- | --- | --- | --- |
|  | |  | |  | | 95% CI | | | |
|  | | Comparison | | CCC estimate | | Lower | | | Upper |
| Active HD-tDCS | | | | | | | | | |
| Easy | | B vs F | | 0.44 | | -0.01 | | | 0.74 |
|  | | F vs M | | 0.21 | | -0.08 | | | 0.47 |
|  | | B vs M | | 0.37 | | -0.09 | | | 0.70 |
| Medium | | B vs F | | **0.79*** | | 0.37 | | | 0.94 |
|  | | F vs M | | 0.32 | | -0.23 | | | 0.71 |
|  | | B vs M | | 0.37 | | -0.17 | | | 0.74 |
| Hard | | B vs F | | 0.45 | | -0.30 | | | 0.86 |
|  | | F vs M | | 0.61 | | -0.01 | | | 0.89 |
|  | | B vs M | | 0.26 | | -0.46 | | | 0.78 |
| Sham HD-tDCS | | | | | | | | | |
| Easy | | B vs F | | **0.73*** | | 0.30 | | | 0.91 |
|  | | F vs M | | 0.40 | | -0.04 | | | 0.71 |
|  | | B vs M | | **0.62*** | | 0.06 | | | 0.88 |
| Medium | | B vs F | | **0.79*** | | 0.28 | | | 0.95 |
|  | | F vs M | | **0.71*** | | 0.41 | | | 0.88 |
|  | | B vs M | | **0.71*** | | 0.46 | | | 0.86 |
| Hard | | B vs F | | **0.75*** | | 0.17 | | | 0.94 |
|  | | F vs M | | 0.61 | | -0.11 | | | 0.91 |
|  | | B vs M | | 0.13 | | -0.57 | | | 0.72 |
| **Table S2.** Biological Motion Wilcoxon Results | | | | | | | | | |
|  | **Comparison** | | **Statistic** | | **p** | | **p adjusted** | **Rank Biserial r** | |
| Active HD-tDCS | | | | | | | | | |
| Easy | B vs F | | 5 | | 1.00 | | 1.00 | 0.00 | |
|  | B vs M | | 1 | | 0.20 | | 0.59 | -0.80 | |
|  | F vs M | | 2 | | 0.36 | | 0.72 | -0.60 | |
| Medium | B vs F | | 4.5 | | 0.50 | | 1.00 | -0.40 | |
|  | B vs M | | 3 | | 0.27 | | 0.82 | -0.60 | |
|  | F vs M | | 7 | | 1.00 | | 1.00 | -0.07 | |
| Hard | B vs F | | 8.5 | | 0.75 | | 1.00 | -0.19 | |
|  | B vs M | | 7 | | 1.00 | | 1.00 | -0.07 | |
|  | F vs M | | 10 | | 1.00 | | 1.00 | -0.05 | |
| Sham HD-tDCS | | | | | | | | | |
| Easy | B vs F | | 12 | | 0.28 | | 0.83 | 0.60 | |
|  | B vs M | | 8 | | 1.00 | | 1.00 | 0.07 | |
|  | F vs M | | 9 | | 0.83 | | 1.00 | -0.14 | |
| Medium | B vs F | | 4 | | 0.41 | | 0.84 | -0.47 | |
|  | B vs M | | 3 | | 0.28 | | 0.84 | -0.60 | |
|  | F vs M | | 5 | | 1.00 | | 1.00 | 0.00 | |
| Hard | B vs F | | 9 | | 0.79 | | 1.00 | 0.20 | |
|  | B vs M | | 13.5 | | 0.60 | | 1.00 | 0.29 | |
|  | F vs M | | 11 | | 0.42 | | 1.00 | 0.47 | |

| **Table S3.** GAF, MADRS, and YMRS Results | | | | | |
| --- | --- | --- | --- | --- | --- |
|  | **Comparison** | **Statistic** | **p** | **p adjusted** | **Rank Biserial r** |
| **Active HD-tDCS** | | | | | |
| GAF | B vs F | 1.00 | 0.41 | 0.84 | -0.90 |
|  | B vs M | 3.00 | 0.28 | 0.84 | -0.71 |
|  | F vs M | 4.50 | 0.50 | 0.84 | -0.57 |
| MADRS | B vs F | 6.00 | 0.85 | 1.00 | -0.43 |
|  | B vs M | 5.50 | 0.68 | 1.00 | -0.48 |
|  | F vs M | 4.50 | 0.50 | 1.00 | -0.57 |
| YMRS | B vs F | 13.00 | 0.18 | 0.53 | 0.24 |
|  | B vs M | 10.50 | 0.50 | 0.72 | 0.00 |
|  | F vs M | 2.00 | 0.36 | 0.72 | -0.81 |
| **Sham HD-tDCS** | | | | | |
| GAF | B vs F | 10.00 | 0.59 | 1.00 | -0.05 |
|  | B vs M | 11.50 | 0.92 | 1.00 | 0.10 |
|  | F vs M | 8.00 | 1.00 | 1.00 | -0.24 |
| MADRS | B vs F | 4.50 | 0.50 | 1.00 | -0.57 |
|  | B vs M | 8.00 | 0.67 | 1.00 | -0.24 |
|  | F vs M | 8.00 | 1.00 | 1.00 | -0.24 |
| YMRS | B vs F | 3.00 | 0.28 | 0.56 | -0.71 |
|  | B vs M | 0.00 | **0.04*** | 0.11 | -1.00 |
|  | F vs M | 11.00 | 1.00 | 1.00 | 0.05 |
| *Note.* Results are shown for the Global Assessment of Function (GAF), Montgomery Asberg Depression Rating Scale (MADRS), and Young Mania Rating Scale (YMRS) between baseline, 5-day, and 1-month timepoints (B/F/M). | | | | | |

**BAC Subdomains**

The BAC App is comprised of 6 tasks indexing working memory (verbal memory, digit sequencing), psychomotor and visual processing speed (token motor), lexical access (verbal fluency), attention and processing speed (symbol coding), and executive functioning (Tower of London). The verbal memory task uses different sets of words at each session as the BAC App was administered at multiple time points.

| **Table S4.** Brief Assessment of Cognition (BAC) Results | | | | | |
| --- | --- | --- | --- | --- | --- |
|  | **Comparison** | **Stat** | **p** | **p adjusted** | **Rank Biserial r** |
| **Active HD-tDCS** | | | | | |
| BAC Composite | B vs F | 14.00 | 0.53 | 1.00 | 0.33 |
|  | B vs M | 11.00 | 1.00 | 1.00 | 0.05 |
|  | F vs M | 6.00 | 0.40 | 1.00 | -0.43 |
| Verbal Memory | B vs F | 11.00 | 0.42 | 0.88 | 0.05 |
|  | B vs M | 9.00 | 0.79 | 0.88 | -0.14 |
|  | F vs M | 5.00 | 0.29 | 0.88 | -0.52 |
| Digit Sequencing | B vs F | 7.00 | 0.58 | 1.00 | -0.33 |
|  | B vs M | 6.00 | 0.79 | 1.00 | -0.43 |
|  | F vs M | 6.00 | 0.79 | 1.00 | -0.43 |
| Token Motor | B vs F | 10.00 | 0.10 | 0.30 | -0.05 |
|  | B vs M | 2.00 | 0.79 | 0.79 | -0.81 |
|  | F vs M | 1.00 | 0.11 | 0.30 | -0.90 |
| Verbal Fluency | B vs F | 11.00 | 1.00 | 1.00 | 0.05 |
|  | B vs M | 5.00 | 0.29 | 0.88 | -0.52 |
|  | F vs M | 7.00 | 0.53 | 1.00 | -0.33 |
| Symbol Coding | B vs F | 12.00 | 0.83 | 1.00 | 0.14 |
|  | B vs M | 14.00 | 0.53 | 1.00 | 0.33 |
|  | F vs M | 14.00 | 0.53 | 1.00 | 0.33 |
| Tower of London | B vs F | 3.00 | 0.58 | 1.00 | -0.71 |
|  | B vs M | 1.00 | 0.20 | 0.60 | -0.90 |
|  | F vs M | 3.00 | 1.00 | 1.00 | -0.71 |
| **Sham HD-tDCS** | | | | | |
| BAC Composite | B vs F | 17.00 | 0.21 | 0.28 | 0.62 |
|  | B vs M | 21.00 | **0.04*** | 0.11 | 1.00 |
|  | F vs M | 18.00 | 0.14 | 0.28 | 0.71 |
| Verbal Memory | B vs F | 16.00 | 0.29 | 0.88 | 0.52 |
|  | B vs M | 15.00 | 0.40 | 0.88 | 0.43 |
|  | F vs M | 5.00 | 0.59 | 0.88 | -0.52 |
| Digit Sequencing | B vs F | 13.00 | 0.67 | 1.00 | 0.24 |
|  | B vs M | 7.00 | 1.00 | 1.00 | -0.33 |
|  | F vs M | 9.00 | 0.83 | 1.00 | -0.14 |
| Token Motor | B vs F | 3.00 | 0.28 | 0.84 | -0.71 |
|  | B vs M | 7.50 | 0.60 | 1.00 | -0.29 |
|  | F vs M | 10.00 | 1.00 | 1.00 | -0.05 |
| Verbal Fluency | B vs F | 9.00 | 0.79 | 1.00 | -0.14 |
|  | B vs M | 14.00 | 0.53 | 1.00 | 0.33 |
|  | F vs M | 14.00 | 0.53 | 1.00 | 0.33 |
| Symbol Coding | B vs F | 6.00 | 0.86 | 1.00 | -0.43 |
|  | B vs M | 14.00 | 0.53 | 1.00 | 0.33 |
|  | F vs M | 14.00 | 0.53 | 1.00 | 0.33 |
| Tower of London | B vs F | 9.00 | 0.20 | 0.60 | -0.14 |
|  | B vs M | 15.00 | 0.40 | 0.80 | 0.43 |
|  | F vs M | 10.00 | 1.00 | 1.00 | -0.05 |

| **Table S5.** Symptom Checklist 90 (SCL-90) Results | | | | | |
| --- | --- | --- | --- | --- | --- |
|  | **Comparison** | **Stat** | ***p*** | ***p* adjusted** | **Rank Biserial *r*** |
| **Active HD-tDCS** | | | | | |
| SCL-90 Total | B vs F | 20.00 | **0.06*** | 0.18 | 0.90 |
|  | B vs M | 10.00 | 1.00 | 1.00 | -0.05 |
|  | F vs M | 1.00 | **0.06*** | 0.18 | -0.90 |
| Somatization | B vs F | 15.00 | **0.06*** | 0.18 | 0.43 |
|  | B vs M | 3.00 | 0.58 | 0.58 | -0.71 |
|  | F vs M | 0.00 | **0.10*** | 0.20 | -1.00 |
| OCD | B vs F | 19.00 | **0.09*** | 0.27 | 0.81 |
|  | B vs M | 12.50 | 0.75 | 0.75 | 0.19 |
|  | F vs M | 3.00 | 0.28 | 0.56 | -0.71 |
| Interpersonal | B vs F | 12.00 | 0.28 | 0.56 | 0.14 |
|  | B vs M | 8.50 | 0.89 | 0.89 | -0.19 |
|  | F vs M | 2.00 | **0.09*** | 0.27 | -0.81 |
| Depression | B vs F | 10.00 | 0.59 | 1.00 | -0.05 |
|  | B vs M | 8.50 | 0.89 | 1.00 | -0.19 |
|  | F vs M | 5.00 | 0.59 | 1.00 | -0.52 |
| Anxiety | B vs F | 21.00 | **0.04*** | 0.11 | 1.00 |
|  | B vs M | 10.00 | 0.59 | 0.59 | -0.05 |
|  | F vs M | 1.00 | 0.20 | 0.40 | -0.90 |
| Phobic | B vs F | 10.00 | **0.10*** | 0.29 | -0.05 |
|  | B vs M | 6.00 | 0.85 | 1.00 | -0.43 |
|  | F vs M | 1.50 | 0.59 | 1.00 | -0.86 |
| Paranoid | B vs F | 10.00 | 0.59 | 1.00 | -0.05 |
|  | B vs M | 7.50 | 1.00 | 1.00 | -0.29 |
|  | F vs M | 3.00 | 0.58 | 1.00 | -0.71 |
| Psychoticism | B vs F | 8.00 | 0.36 | 0.72 | -0.24 |
|  | B vs M | 6.00 | 0.18 | 0.54 | -0.43 |
|  | F vs M | 1.00 | 1.00 | 1.00 | -0.90 |
| **Sham HD-tDCS** | | | | | |
| SCL-90 Total | B vs F | 1.00 | **0.06*** | 0.18 | -0.90 |
|  | B vs M | 5.00 | 0.29 | 0.59 | -0.52 |
|  | F vs M | 5.00 | 0.29 | 0.59 | -0.52 |
| Somatization | B vs F | 8.00 | 0.67 | 0.67 | -0.24 |
|  | B vs M | 2.00 | **0.09*** | 0.19 | -0.81 |
|  | F vs M | 0.00 | **0.04*** | 0.11 | -1.00 |
| OCD | B vs F | 7.50 | 0.60 | 1.00 | -0.29 |
|  | B vs M | 10.00 | 1.00 | 1.00 | -0.05 |
|  | F vs M | 12.00 | 0.83 | 1.00 | 0.14 |
| Interpersonal | B vs F | 1.00 | 0.10 | 0.31 | -0.90 |
|  | B vs M | 6.00 | 0.40 | 0.80 | -0.43 |
|  | F vs M | 9.00 | 0.83 | 0.83 | -0.14 |
| Depression | B vs F | 5.50 | 0.68 | 1.00 | -0.48 |
|  | B vs M | 8.00 | 0.67 | 1.00 | -0.24 |
|  | F vs M | 6.00 | 0.40 | 1.00 | -0.43 |
| Anxiety | B vs F | 4.00 | 0.41 | 0.50 | -0.62 |
|  | B vs M | 2.50 | 0.11 | 0.34 | -0.76 |
|  | F vs M | 4.50 | 0.25 | 0.50 | -0.57 |
| Phobic | B vs F | 8.00 | 0.67 | 0.67 | -0.24 |
|  | B vs M | 3.00 | 0.14 | 0.43 | -0.71 |
|  | F vs M | 2.00 | 0.18 | 0.43 | -0.81 |
| Paranoid | B vs F | 1.50 | 0.14 | 0.41 | -0.86 |
|  | B vs M | 6.00 | 0.40 | 0.80 | -0.43 |
|  | F vs M | 8.50 | 0.75 | 0.80 | -0.19 |
| Psychoticism | B vs F | 9.00 | 0.83 | 1.00 | -0.14 |
|  | B vs M | 10.50 | 1.00 | 1.00 | 0.00 |
|  | F vs M | 14.00 | 0.53 | 1.00 | 0.33 |

**Imputation**

Average missingness consisted of 0% at baseline, 4.75% at day 5, 14.04% at 1 month (Average overall missingness 8.03%). Core predictors included in the regression models for imputation of missing values included the individual, timepoint, and stimulation condition as fixed effects. Ten iterations were run in the model. Density plots for missing values were visually inspected to ensure good fit of predicted values. Data was combined to produce a mean value obtained across the imputed datasets for missing values.

| **Table S6.** Unimputed PANSS Results | | | | | |
| --- | --- | --- | --- | --- | --- |
|  | **Comparison** | **Statistic** | **p** | **p adjusted** | **Rank Biserial r** |
| Active HD-tDCS | | | | | |
| Total | B vs F | 20 | **0.03*** | **0.09*** | 0.90 |
|  | B vs M | 14 | **0.05*** | 0.11 | 0.87 |
|  | F vs M | 7 | 0.29 | 0.29 | 0.40 |
| Positive | B vs F | 6 | 0.09 | 0.27 | 1.00 |
|  | B vs M | 11 | 0.21 | 0.42 | 0.47 |
|  | F vs M | 5.5 | 0.50 | 0.50 | 0.10 |
| Negative | B vs F | 6.5 | 0.36 | 0.71 | 0.30 |
|  | B vs M | 8.5 | 0.13 | 0.40 | 0.70 |
|  | F vs M | 3 | 0.61 | 0.71 | 0.00 |
| General | B vs F | 14.5 | 0.23 | 0.46 | 0.38 |
|  | B vs M | 17.5 | **0.09*** | 0.26 | 0.67 |
|  | F vs M | 9 | 0.39 | 0.46 | 0.20 |
| Sham HD-tDCS | | | | | |
| Total | B vs F | 8 | 0.74 | 1.00 | -0.24 |
|  | B vs M | 4.5 | 0.83 | 1.00 | -0.40 |
|  | F vs M | 1 | 0.95 | 1.00 | -0.80 |
| Positive | B vs F | 8.5 | 0.13 | 0.40 | 0.70 |
|  | B vs M | 8.5 | 0.45 | 0.89 | 0.13 |
|  | F vs M | 1 | 0.95 | 0.95 | -0.80 |
| Negative | B vs F | 11 | 0.50 | 1.00 | 0.05 |
|  | B vs M | 6 | 0.43 | 1.00 | 0.20 |
|  | F vs M | 5 | 0.57 | 1.00 | 0.00 |
| General | B vs F | 6.5 | 0.83 | 1.00 | -0.38 |
|  | B vs M | 1 | 0.95 | 1.00 | -0.80 |
|  | F vs M | 0 | 0.98 | 1.00 | -1.00 |

| **Table S7.** Unimputed AVI EEG Results | | | | | |
| --- | --- | --- | --- | --- | --- |
|  | **Comparison** | **Statistic** | **p** | **p adjusted** | **Rank Biserial r** |
| Active HD-tDCS | | | | | |
| ASSR | B vs F | 9 | 0.79 | 0.79 | 0.20 |
|  | B vs M | 2 | 0.18 | 0.36 | -0.73 |
|  | F vs M | 0 | **0.06*** | 0.18 | -1.00 |
| AVI | B vs F | 15 | **0.06*** | 0.18 | 1.00 |
|  | B vs M | 13 | 0.18 | 0.36 | 0.73 |
|  | F vs M | 9 | 0.79 | 0.79 | 0.20 |
| VSSR | B vs F | 2 | 0.18 | 0.36 | -0.73 |
|  | B vs M | 1 | 0.11 | 0.32 | -0.87 |
|  | F vs M | 6 | 0.79 | 0.79 | -0.20 |
| Sham HD-tDCS | | | | | |
| ASSR | B vs F | 6 | 0.79 | 1.00 | -0.20 |
|  | B vs M | 6 | 0.18 | 0.54 | 1.00 |
|  | F vs M | 7 | 0.58 | 1.00 | 0.40 |
| AVI | B vs F | 11 | 0.42 | 1.00 | 0.47 |
|  | B vs M | 4 | 0.79 | 1.00 | 0.33 |
|  | F vs M | 7 | 0.58 | 1.00 | 0.40 |
| VSSR | B vs F | 5 | 0.59 | 1.00 | -0.33 |
|  | B vs M | 5 | 0.42 | 1.00 | 0.67 |
|  | F vs M | 5 | 1.00 | 1.00 | 0.00 |

| **Table S8.** Unimputed rsEEG Results | | | | | |
| --- | --- | --- | --- | --- | --- |
|  | **Comparison** | **Stat** | ***p*** | ***p adjusted*** | **Rank Biserial *r*** |
| **Active HD-tDCS** | | | | | |
| Delta/Theta | B vs F | 10.00 | 1.00 | 1.00 | -0.05 |
|  | B vs M | 3.00 | 0.14 | 0.43 | -0.71 |
|  | F vs M | 7.00 | 0.53 | 1.00 | -0.33 |
| ROI Delta/Theta | B vs F | 14.00 | 0.53 | 1.00 | 0.33 |
|  | B vs M | 7.00 | 0.53 | 1.00 | -0.33 |
|  | F vs M | 5.00 | 0.29 | 0.88 | -0.52 |
| Alpha | B vs F | 8.00 | 0.67 | 0.88 | -0.24 |
|  | B vs M | 15.00 | 0.40 | 0.88 | 0.43 |
|  | F vs M | 16.00 | 0.29 | 0.88 | 0.52 |
| ROI Alpha | B vs F | 10.00 | 1.00 | 1.00 | -0.05 |
|  | B vs M | 20.00 | **0.06*** | 0.18 | 0.90 |
|  | F vs M | 14.00 | 0.53 | 1.00 | 0.33 |
| Beta | B vs F | 14.00 | 0.53 | 0.88 | 0.33 |
|  | B vs M | 15.00 | 0.40 | 0.88 | 0.43 |
|  | F vs M | 16.00 | 0.29 | 0.88 | 0.52 |
| ROI Beta | B vs F | 6.00 | 0.40 | 0.88 | -0.43 |
|  | B vs M | 13.00 | 0.67 | 0.88 | 0.24 |
|  | F vs M | 16.00 | 0.29 | 0.88 | 0.52 |
| Gamma | B vs F | 11.00 | 1.00 | 1.00 | 0.05 |
|  | B vs M | 14.00 | 0.53 | 1.00 | 0.33 |
|  | F vs M | 16.00 | 0.29 | 0.88 | 0.52 |
| ROI Gamma | B vs F | 6.00 | 0.40 | 1.00 | -0.43 |
|  | B vs M | 9.00 | 0.83 | 1.00 | -0.14 |
|  | F vs M | 15.00 | 0.40 | 1.00 | 0.43 |
| **Sham HD-tDCS** | | | | | |
| Delta/Theta | B vs F | 12.00 | 0.83 | 1.00 | 0.14 |
|  | B vs M | 7.00 | 0.58 | 1.00 | 0.40 |
|  | F vs M | 7.00 | 0.58 | 1.00 | 0.40 |
| ROI Delta/Theta | B vs F | 11.00 | 1.00 | 1.00 | 0.05 |
|  | B vs M | 7.00 | 0.58 | 1.00 | 0.40 |
|  | F vs M | 6.00 | 0.86 | 1.00 | 0.20 |
| Alpha | B vs F | 4.00 | 0.21 | 0.60 | -0.62 |
|  | B vs M | 6.00 | 0.86 | 0.86 | 0.20 |
|  | F vs M | 9.00 | 0.20 | 0.60 | 0.80 |
| ROI Alpha | B vs F | 10.00 | 1.00 | 1.00 | -0.05 |
|  | B vs M | 5.00 | 1.00 | 1.00 | 0.00 |
|  | F vs M | 7.00 | 0.58 | 1.00 | 0.40 |
| Beta | B vs F | 12.00 | 0.83 | 1.00 | 0.14 |
|  | B vs M | 5.00 | 1.00 | 1.00 | 0.00 |
|  | F vs M | 6.00 | 0.86 | 1.00 | 0.20 |
| ROI Beta | B vs F | 11.00 | 1.00 | 1.00 | 0.05 |
|  | B vs M | 5.00 | 1.00 | 1.00 | 0.00 |
|  | F vs M | 7.00 | 0.58 | 1.00 | 0.40 |
| Gamma | B vs F | 13.00 | 0.67 | 1.00 | 0.24 |
|  | B vs M | 5.00 | 1.00 | 1.00 | 0.00 |
|  | F vs M | 4.00 | 0.86 | 1.00 | -0.20 |
| ROI Gamma | B vs F | 12.00 | 0.83 | 1.00 | 0.14 |
|  | B vs M | 4.00 | 0.86 | 1.00 | -0.20 |
|  | F vs M | 6.00 | 0.86 | 1.00 | 0.20 |

| **Table S9.** Unimputed Biological Motion Wilcoxon Results | | | | | |
| --- | --- | --- | --- | --- | --- |
|  | **Comparison** | **Stat** | **p** | **p adjusted** | **Rank Biserial r** |
| **Active HD-tDCS** | | | | | |
| Easy | B vs F | 3 | 1.00 | 1.00 | -0.60 |
|  | B vs M | 0 | 0.17 | 0.52 | -1.00 |
|  | F vs M | 1 | 0.42 | 0.85 | -0.87 |
| Medium | B vs F | 3.5 | 0.71 | 1.00 | -0.53 |
|  | B vs M | 2 | 0.34 | 1.00 | -0.73 |
|  | F vs M | 6 | 0.85 | 1.00 | -0.20 |
| Hard | B vs F | 6.5 | 0.89 | 1.00 | -0.13 |
|  | B vs M | 4 | 0.86 | 1.00 | -0.47 |
|  | F vs M | 6 | 0.79 | 1.00 | -0.20 |
| **Sham HD-tDCS** | | | | | |
| Easy | B vs F | 12 | 0.28 | 0.55 | 0.14 |
|  | B vs M | 1 | 1.00 | 1.00 | -0.67 |
|  | F vs M | 0 | 0.18 | 0.54 | -1.00 |
| Medium | B vs F | 4 | 0.41 | 0.82 | -0.62 |
|  | B vs M | 0 | 0.17 | 0.52 | -1.00 |
|  | F vs M | 1 | 1.00 | 1.00 | -0.67 |
| Hard | B vs F | 9 | 0.79 | 1.00 | -0.14 |
|  | B vs M | 3.5 | 1.00 | 1.00 | 0.17 |
|  | F vs M | 1 | 1.00 | 1.00 | -0.67 |

| **Table S10.** Unimputed GAF, MADRS, and YMRS Results | | | | | |
| --- | --- | --- | --- | --- | --- |
|  | **Comparison** | **Stat** | **p** | **p adjusted** | **Rank Biserial r** |
| **Active HD-tDCS** | | | | | |
| GAF | B vs F | 1.00 | 0.41 | 0.84 | -0.90 |
|  | B vs M | 3.00 | 0.28 | 0.84 | -0.71 |
|  | F vs M | 4.50 | 0.50 | 0.84 | -0.57 |
| MADRS | B vs F | 6.00 | 0.85 | 1.00 | -0.43 |
|  | B vs M | 5.50 | 0.68 | 1.00 | -0.48 |
|  | F vs M | 4.50 | 0.50 | 1.00 | -0.57 |
| YMRS | B vs F | 13.00 | 0.18 | 0.53 | 0.24 |
|  | B vs M | 10.50 | 0.50 | 0.72 | 0.00 |
|  | F vs M | 2.00 | 0.36 | 0.72 | -0.81 |
| **Sham HD-tDCS** | | | | | |
| GAF | B vs F | 10.00 | 0.59 | 1.00 | -0.05 |
|  | B vs M | 5.50 | 0.68 | 1.00 | -0.27 |
|  | F vs M | 3.00 | 0.58 | 1.00 | -0.60 |
| MADRS | B vs F | 4.50 | 0.50 | 1.00 | -0.57 |
|  | B vs M | 5.00 | 0.59 | 1.00 | -0.33 |
|  | F vs M | 5.00 | 1.00 | 1.00 | -0.33 |
| YMRS | B vs F | 3.00 | 0.58 | 0.58 | -0.60 |
|  | B vs M | 0.00 | 0.10 | 0.29 | -1.00 |
|  | F vs M | 0.00 | 0.10 | 0.29 | -1.00 |

| **Table S11.** Unimputed BAC Results | | | | | |
| --- | --- | --- | --- | --- | --- |
|  | **Comparison** | **Stat** | **p** | **p adjusted** | **Rank Biserial r** |
| **Active HD-tDCS** | | | | | |
| BACS Composite | B vs F | 14.00 | 0.53 | 1.00 | 0.33 |
|  | B vs M | 11.00 | 1.00 | 1.00 | 0.05 |
|  | F vs M | 6.00 | 0.40 | 1.00 | -0.43 |
| Verbal Memory | B vs F | 11.00 | 0.42 | 0.88 | 0.05 |
|  | B vs M | 9.00 | 0.79 | 0.88 | -0.14 |
|  | F vs M | 5.00 | 0.29 | 0.88 | -0.52 |
| Digit Sequencing | B vs F | 7.00 | 0.58 | 1.00 | -0.33 |
|  | B vs M | 6.00 | 0.79 | 1.00 | -0.43 |
|  | F vs M | 6.00 | 0.79 | 1.00 | -0.43 |
| Token Motor | B vs F | 10.00 | 0.10 | 0.30 | -0.05 |
|  | B vs M | 2.00 | 0.79 | 0.79 | -0.81 |
|  | F vs M | 1.00 | 0.11 | 0.30 | -0.90 |
| Verbal Fluency | B vs F | 11.00 | 1.00 | 1.00 | 0.05 |
|  | B vs M | 5.00 | 0.29 | 0.88 | -0.52 |
|  | F vs M | 7.00 | 0.53 | 1.00 | -0.33 |
| Symbol Coding | B vs F | 12.00 | 0.83 | 1.00 | 0.14 |
|  | B vs M | 14.00 | 0.53 | 1.00 | 0.33 |
|  | F vs M | 14.00 | 0.53 | 1.00 | 0.33 |
| Tower of London | B vs F | 3.00 | 0.58 | 1.00 | -0.71 |
|  | B vs M | 1.00 | 0.20 | 0.60 | -0.90 |
|  | F vs M | 3.00 | 1.00 | 1.00 | -0.71 |
| **Sham HD-tDCS** | | | | | |
| BACS Composite | B vs F | 17.00 | 0.21 | 0.40 | 0.62 |
|  | B vs M | 10.00 | 0.10 | 0.30 | 1.00 |
|  | F vs M | 9.00 | 0.20 | 0.40 | 0.80 |
| Verbal Memory | B vs F | 16.00 | 0.29 | 0.88 | 0.52 |
|  | B vs M | 7.00 | 0.58 | 1.00 | 0.40 |
|  | F vs M | 3.00 | 1.00 | 1.00 | -0.40 |
| Digit Sequencing | B vs F | 13.00 | 0.67 | 1.00 | 0.24 |
|  | B vs M | 1.00 | 0.42 | 1.00 | -0.80 |
|  | F vs M | 4.00 | 0.86 | 1.00 | -0.20 |
| Token Motor | B vs F | 3.00 | 0.28 | 0.81 | -0.71 |
|  | B vs M | 1.50 | 0.27 | 0.81 | -0.70 |
|  | F vs M | 3.00 | 0.58 | 0.81 | -0.40 |
| Verbal Fluency | B vs F | 9.00 | 0.79 | 0.79 | -0.14 |
|  | B vs M | 8.00 | 0.36 | 0.72 | 0.60 |
|  | F vs M | 9.00 | 0.20 | 0.60 | 0.80 |
| Symbol Coding | B vs F | 6.00 | 0.86 | 1.00 | -0.43 |
|  | B vs M | 6.00 | 0.86 | 1.00 | 0.20 |
|  | F vs M | 8.00 | 0.36 | 1.00 | 0.60 |
| Tower of London | B vs F | 9.00 | 0.20 | 0.60 | -0.14 |
|  | B vs M | 9.00 | 0.20 | 0.60 | 0.80 |
|  | F vs M | 7.00 | 0.58 | 0.60 | 0.40 |

**Power Analyses**

Power analyses were performed for significant within-subject changes in PANSS, ASSR, AVI, rsEEG alpha over the rSTS, and SCL90-R Total. Post-hoc analyses calculated achieved power from one- and two-tailed Wilcoxon tests in this 6-subject pilot study with α=.10. A priori analyses calculated required sample sizes to guide a well-powered follow-up study with ≥80% power for two-tailed Wilcoxon tests with α=.05.

| **Table S12.**  *Power analyses* | | | | | | |
| --- | --- | --- | --- | --- | --- | --- |
|  |  |  | Post hoc | | A priori | |
| Measure | Timepoints | Rank biserial ES | N per group | Achieved power | Required N per group | Actual power (all 2-tailed) |
| PANSS Total | B to F | 0.9 | 6 | 75% | 13 | 83% |
| PANSS Total | B to M | 0.87 | 6 | 73% | 14 | 83% |
| PANSS Pos | B to F | 1 | 6 | 81% | 11 | 83% |
| PANSS Gen | B to M | 0.67 | 6 | 57% | 21 | 81% |
| ASSR | F to M | -1 | 6 | 65% | 11 | 83% |
| AVI | B to F | 1 | 6 | 65% | 11 | 83% |
| rsEEG ROI α | B to M | 0.9 | 6 | 58% | 13 | 83% |
| SCL90R Total | B to F | 0.9 | 6 | 58% | 13 | 83% |
| *Note.* PANSS post hoc analyses performed for one-tailed tests. All others performed for two-tailed tests. | | | | | | |

**Sensation Questionnaire**

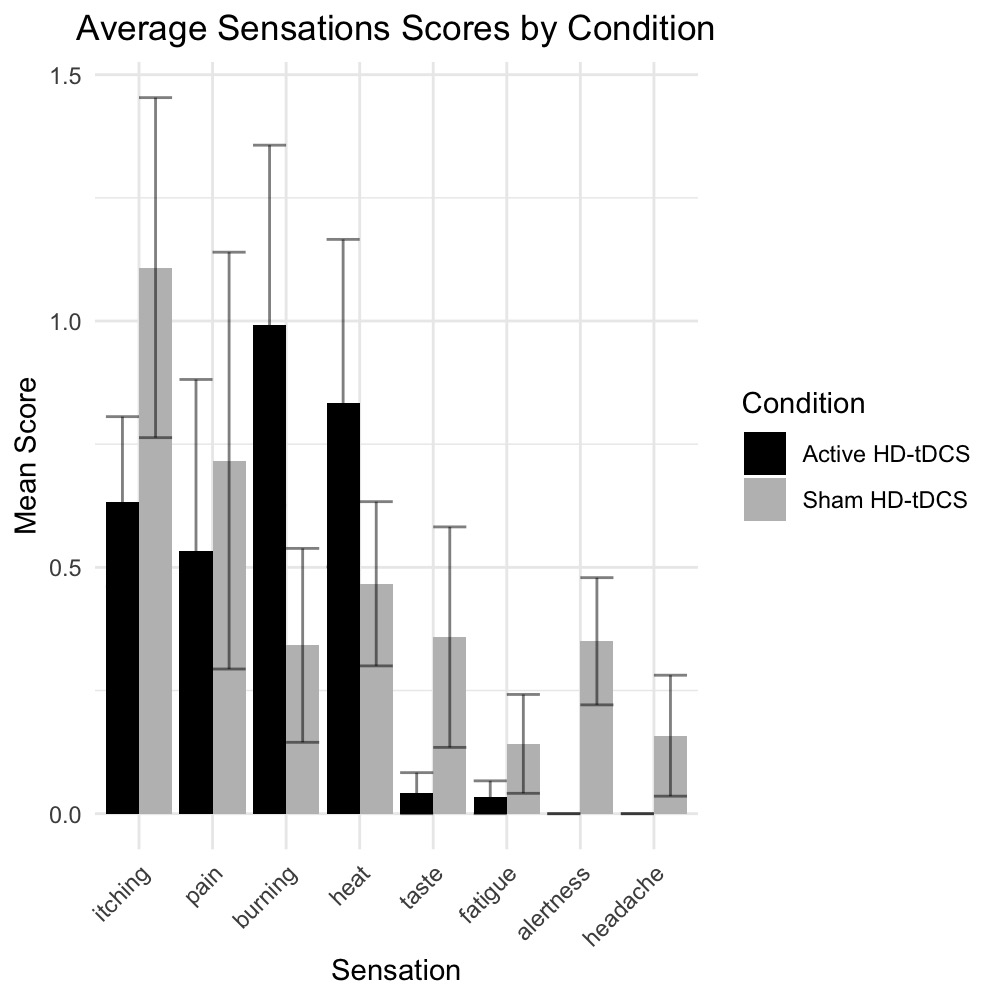

**Figure S3.** Sensation Questionnaire Responses. 0=None, 1=Mild, 2=Moderate, 3=Strong. Bars show mean with standard error for each group.

| **Table S13.** Sensation Questionnaire Statistics | | |
| --- | --- | --- |
| **Sensation Variable** | **W statistic** | **p** |
| Itching | 23 | 0.46 |
| Pain | 19.5 | 0.86 |
| Burning | 11 | 0.28 |
| Heat | 12.5 | 0.41 |
| Taste | 25 | 0.21 |
| Fatigue | 22 | 0.46 |
| Alertness | 30 | **0.03*** |
| Headache | 24 | 0.18 |
| *Note.* Statistics test differences between groups | | |
